# Prevalence of cognitive impairment in older adult patients with type 2 diabetes mellitus in the family medicine outpatient clinic of HGR C/MF No.1, Cuernavaca, Morelos

**DOI:** 10.64898/2025.12.02.25341516

**Authors:** Julio Cesar Flores Albarran, Iris García Orihuela, Andrea Marai Albarran Riojas

**Author notes:** **Corresponding Author:** Julio Cesar Flores Albarran.

## Abstract

**Background:** Type 2 diabetes mellitus is a heterogeneous metabolic disorder characterized by hyperglycemia due to impaired insulin secretion, defective insulin action, or both. It is the main type of diabetes in older adults and the second leading cause of death in the Mexican population. An increased prevalence of cognitive impairment has been found in older adults living with this pathology, and it is estimated that up to 50% of people suffering from it are not diagnosed by primary care physicians.

**Objective:** To determine the prevalence of cognitive impairment in older adults with type 2 diabetes mellitus treated at the family medicine outpatient clinic of HGR C/MF No.1.

**Methods:** Study design: Cross-sectional, descriptive, observational. Study population: Older adult patients with type 2 diabetes mellitus assigned to the family medicine outpatient clinic of HGR C/MF No.1. Data were collected from medical notes in the SIMF platform according to established criteria. Sociodemographic data were recorded, and the Mini-Mental State Examination was applied to determine the degree of cognitive impairment. Subsequently, data were entered into Excel for cleaning and further analysis in a statistical program.

**Results:** Of 241 patients, 80.1% presented no cognitive impairment, 16.6% showed mild impairment, and 3.3% moderate impairment. Women (67.2%), age 60-70 years, and low schooling predominated.

**Conclusions:** The majority did not present impairment; mild and moderate cases were associated with female sex, low educational level, longer duration of evolution, and family history of dementia.

## INTRODUCTION

Type 2 Diabetes Mellitus (T2DM) is a heterogeneous metabolic disorder characterized by hyperglycemia due to impaired insulin secretion, defective insulin action, or both. According to the American Diabetes Association (ADA), T2DM is the main type of diabetes present in older adults.

Globally, the prevalence of T2DM has risen dramatically. In 2014, approximately 422 million adults lived with diabetes, and this figure is expected to reach 642 million by 2040. In Mexico, the 2022 National Health and Nutrition Survey (ENSANUT) reported a total diabetes prevalence of 18.3% in the general population, rising to 37% in the age group of 60 years and older.

Pathophysiologically, insulin deficiency and resistance lead to hyperglycemia. In the brain, insulin receptors in the hippocampus play a key role in memory preservation. Metabolic dysfunction in diabetes, including endothelial dysfunction and chronic micro-inflammation, has been linked to cognitive decline and structural changes in the brain, such as white matter alterations.

Cognitive impairment is a clinical syndrome characterized by the loss or deterioration of mental functions across various domains, including memory, orientation, and judgment. It is estimated that 5% to 8% of the global population over 60 years of age suffers from cognitive impairment. In Mexico, the prevalence is reported around 8% in those over 65. However, in patients with T2DM, the risk of cognitive impairment is nearly doubled. Despite this, cognitive health is not systematically evaluated in primary care, and it is estimated that up to 50% of patients with cognitive impairment remain undiagnosed.

The coexistence of T2DM and cognitive impairment in older adults complicates clinical management. Therefore, this study aims to determine the prevalence of cognitive impairment in older adults with T2DM treated at the family medicine outpatient clinic of HGR C/MF No.1 in Cuernavaca, Morelos, to provide local data that can improve early detection and comprehensive care strategies.

## MATERIALS AND METHODS

### Study Design and Setting

A cross-sectional, descriptive, and observational study was conducted at the Family Medicine Outpatient Clinic of the General Regional Hospital with Family Medicine (HGR C/MF) No. 1, belonging to the Mexican Institute of Social Security (IMSS) in Cuernavaca, Morelos. The data collection period took place from November 1, 2024, to January 31, 2025.

### Participants and Sample Size

The study population consisted of older adults (aged 60 years or older) with a confirmed diagnosis of Type 2 Diabetes Mellitus (T2DM). The sample size was calculated using the formula for finite populations, based on a total universe of 8,856 patients, a 95% confidence interval, and a 6% margin of error, resulting in a sample of 241 participants. A non-probabilistic convenience sampling method was used to select patients attending their regular medical appointments.

### Eligibility Criteria

- **Inclusion:** Patients >60 years old with T2DM diagnosis assigned to the clinic who agreed to participate and signed the informed consent.
- **Exclusion:** Patients with a history of neurological or psychiatric diseases diagnosed prior to T2DM, patients with disabilities preventing communication, and those currently using psychotropic drugs.
- **Elimination:** Participants who decided to withdraw from the survey or presented incomplete questionnaires.

### Data Collection and Instruments

Data were collected directly during the consultation. A data collection instrument was designed in two sections:

#### 1. Sociodemographic and Clinical Profile

Age, sex, schooling level, time since T2DM diagnosis, and family history of dementia.

#### 2. Cognitive Assessment

The Mini-Mental State Examination (MMSE) was administered. This instrument has been validated for the Mexican population with a sensitivity of 82% and specificity of 84% (Cronbach’s alpha 0.91).

- *Scoring Adjustment:* For patients with 0-4 years of schooling, questions 14 to 19 were excluded, and 8 points were added to the result to control for educational bias.
- *Cut-off points:* 24-30 points (Normal), 19-23 points (Mild impairment), 14-18 points (Moderate impairment), and <14 points (Severe impairment).

### Statistical Analysis

Data were entered into a Microsoft Excel spreadsheet for validation and cleaning. Statistical analysis was performed using Stata version 11.1. Descriptive statistics were used, reporting categorical variables (sex, schooling, impairment grade) as absolute frequencies and percentages.

### Ethical Considerations

The study protocol was reviewed and approved by the Local Health Research Committee 1701 of the IMSS (Registration Number: R-2024-1701-058). The research adhered to the principles of the Declaration of Helsinki and the General Health Law regarding Health Research. Written informed consent was obtained from all participants. The study was classified as minimal risk.

## RESULTS

A total of 241 older adult patients with a previous diagnosis of Type 2 Diabetes Mellitus were included in the study. The study population was predominantly female (n=162; 67.2%). Regarding age, the largest group was 60 to 70 years old (n=147; 61.0%), followed by 71 to 80 years (n=70; 29.0%).

Regarding educational attainment, 36.9% had primary education, while 17.0% had no formal schooling. Only 0.8% had postgraduate studies. The time since T2DM diagnosis was evenly distributed among <10 years (32%), 10-19 years (31.5%), and 20-30 years (32%). A family history of dementia was reported by 15.4% of participants.

### Prevalence of Cognitive Impairment

According to the Mini-Mental State Examination (MMSE) scores, 193 patients (80.1%) showed no cognitive impairment. Cognitive impairment was identified in 48 patients (19.9%), distributed as follows: 40 (16.6%) with mild impairment and 8 (3.3%) with moderate impairment. No cases of severe impairment were found.

### Factors Associated with Cognitive Impairment

- **Sex:** In cases of mild and moderate impairment, the female sex predominated (82.5% and 75.0%, respectively).
- **Age:** The 60-70 age group concentrated the highest number of impairment cases (23 mild, 5 moderate), although the proportion of impairment was slightly higher in the 71-80 group relative to its size.
- **Education:** Mild impairment was most frequent in patients with primary education (47.5%) and no schooling (22.5%). Patients with higher education levels (undergraduate/postgraduate) mostly remained without deterioration.
- **Duration of T2DM:** Moderate impairment was most frequent in patients with 20-30 years of diagnosis (50.0%).

**Table 1.** Sociodemographic and Clinical Characteristics of the Study Population (N=241)

| Variable | Category | n | % |
| --- | --- | --- | --- |
| Sex | Female | 162 | 67.2% |
|  | Male | 79 | 32.8% |
| Age Group | 60 - 70 years | 147 | 61.0% |
|  | 71 - 80 years | 70 | 29.0% |

| **Variable** | **Category** | **n** | **%** |
| --- | --- | --- | --- |
|  | > 80 years | 24 | 10.0% |
| **Schooling** | No schooling | 41 | 17.0% |
|  | Primary | 89 | 36.9% |
|  | Secondary | 42 | 17.4% |
|  | High School / Technical | 39 | 16.2% |
|  | Undergraduate | 28 | 11.6% |
|  | Postgraduate | 2 | 0.8% |
| **Time since T2DM Dx** | < 10 years | 77 | 32.0% |
|  | 10 - 19 years | 76 | 31.5% |
|  | 20 - 30 years | 77 | 32.0% |
|  | > 30 years | 11 | 4.5% |
| **Family History of Dementia** | Yes | 37 | 15.4% |
|  | No | 204 | 84.6% |

**Table 2.** Prevalence of Cognitive Impairment (MMSE)

| **Grade of Impairment** | **n** | **%** |
| --- | --- | --- |
| **No Impairment** | 193 | 80.1% |
| **Mild Impairment** | 40 | 16.6% |
| **Moderate Impairment** | 8 | 3.3% |
| **Severe Impairment** | 0 | 0.0% |
| **Total** | **241** | **100%** |

## DISCUSSION

The results of this study show that the majority of older adults with type 2 diabetes mellitus evaluated in the family medicine outpatient clinic maintained a normal cognitive state (80.1%), while 19.9% presented some degree of cognitive impairment, mainly mild (16.6%) and to a lesser extent moderate (3.3%)^1^. This finding, although lower than the 36.7% prevalence reported by Ramos-Domínguez et al. (2020) in a similar Mexican population, highlights the presence of a subgroup at risk requiring priority clinical attention^2^.

Advanced age was a relevant factor; although the highest absolute number of cases was in the 60-70 age group, the proportion of impairment was higher in the 71-80 group^3^. This is consistent with Murata et al. (2015), who identified age as a main determinant of cognitive decline in metabolic diseases^4^. Education also played a crucial role: patients with low educational levels (primary or no schooling) concentrated most cases of impairment, supporting the “cognitive reserve” theory described by Gao-Yuxia (2016)^5^.

Regarding the duration of T2DM, moderate impairment was most frequent in patients with 20 to 30 years of diagnosis (50.0%)^6^. This aligns with González-Bárcenas et al. (2019), who suggest that prolonged exposure to hyperglycemia favors progressive neurodegeneration^7^.

## Limitations

The main limitation of this work is its cross-sectional design, which does not allow establishing causal relationships. Additionally, the sample was limited to a single medical unit, and variables such as glycemic control (HbA1c) or depression were not included in the analysis^8^.

## CONCLUSIONS

1. The prevalence of cognitive impairment in the study population was 19.9% (16.6% mild and 3.3% moderate). The hypothesis of a prevalence $\ge$36.7% was not confirmed^9999^.
2. Low educational levels were strongly associated with the presence of cognitive impairment, whereas patients with higher education remained mostly without deterioration^10^.
3. Patients with a T2DM duration of 20 to 30 years showed the highest proportion of moderate impairment^11^.
4. These results underscore the importance of routine cognitive assessment in older adults with diabetes, especially those with identified risk factors such as low schooling and long-standing disease^12^.

## Data Availability

All data produced in the present study are available upon reasonable request to the authors.

## Notes

### Competing Interest Statement

The authors have declared no competing interest.

### Funding Statement

This study was self-funded by the authors and did not receive any specific grant from funding agencies in the public, commercial, or not-for-profit sectors.

### Author Declarations

The Comite Local de Investigacion en Salud 1701 of the Instituto Mexicano del Seguro Social, Hospital General Regional con Medicina Familiar No. 1 gave ethical approval for this work.

